# A meta-analysis of genome-wide association studies in 614,243 individuals identifies 59 novel susceptibility loci underlying Dupuytren’s contracture

**DOI:** 10.1101/2022.06.07.22276119

**Authors:** Fedik Rahimov, Jacob F. Degner, John S. Lee, Xiuwen Zheng, FinnGen, Jozsef Karman, Howard J. Jacob, Jeffrey F. Waring

## Abstract

Dupuytren’s contracture or disease (DD) is a disabling, fibroproliferative disease of the hand that affects up to 25% of people of northwestern European descent. It typically manifests in adulthood and many affected individuals have a positive family history, yet the genetic architecture of DD is not completely understood. We conducted genome-wide association studies (GWAS) of DD in 8,846 cases and 347,659 population controls from the UK Biobank resource, and 4,616 cases and 253,122 population controls from the FinnGen study. We combined these datasets with a meta-analysis, which represents the largest GWAS conducted in DD to date including 13,462 cases. We identified 83 loci with genome-wide significance of p < 5 × 10^-8^. We replicated association at the 24 previously reported loci and discovered 59 novel loci, substantially increasing the number of risk loci for DD. Colocalization with expression quantitative trait loci and overlap with genes linked to phenotypically matched human Mendelian disorders or animal models support causal roles for at least 30 genes with high confidence. Among these, fifteen genes cause rare limb abnormalities when mutated, an observation that may potentially shed light on hand specificity of DD phenotype. Gene enrichment analysis revealed predominant role of connective tissue development and maintenance and extracellular matrix homeostasis but limited or no role of inflammatory processes in disease causality. These findings provide key insights into the biological mechanisms underlying DD and identify genetically informed therapeutic targets for DD and possibly for other fibrotic diseases.

## Introduction

Dupuytren’s disease (DD [MIM: 126900]), also known as palmar fascial fibromatosis, is a chronic and progressive condition characterized by contraction of the flesh under the skin of the palm that typically leads to permanently bent fingers. DD is caused by an excessive proliferation of contractile, fibroblastic cells called myofibroblasts, within the palmar fascia of the hand. Myofibroblasts derived from DD nodules characteristically express high levels of the alpha-smooth muscle actin (α-SMA) protein and secrete collagen proteins, predominantly of types I and III, into the extracellular matrix (ECM). In addition, affected areas often display inflammation.^1^ However, the underlying causes that trigger fibroblast activation or that drive persistent hyperproliferation of myofibroblasts are poorly understood. It is thought that external factors, for instance those involved in manual labor occupations that expose hands to frequent mechanical vibrations,^2^ may trigger disease development in genetically predisposed individuals.

The prevalence of DD varies significantly depending on age, sex, and ethnic background.^3,4^ In general, DD is more common in advancing age – in particular, in men of northwestern European origin. For example, in the UK the annual incidence rate of new cases among men was estimated at 34.3 per 100,000 men between 1991 and 1992.^5^ The number of people diagnosed with DD also increases with age, with the estimated mean prevalence rising from 12% to 29% between ages 55 and 75 years.^4^ Furthermore, men typically present symptoms on average 10 years earlier (mean age 55 years) than women.^6^

Fibrotic diseases are a major cause of morbidity and mortality with a significant burden on public healthcare.^7^ Hand impairment in DD can affect the quality of life, however, DD can surgically be corrected and by itself is not associated with significant mortality or morbidity. We hypothesize that due to the key role of myofibroblasts in disease pathogenesis, DD is a good human model disease to investigate the underlying genetic factors that predispose to increased tissue fibrosis as myofibroblasts are thought to play a central role in essentially all fibrotic diseases such as systemic sclerosis and idiopathic pulmonary fibrosis (IPF). Therefore, gene products that play a role in Dupuytren’s fibrosis can potentially be explored as novel therapeutic targets for other fibrotic diseases.

Although DD is a complex disorder with both genetic and environmental risk factors contributing to disease etiology, DD has a strong heritable component with an estimated overall heritability of 80% from a large twin study in Denmark.^8^ In addition, high sibling recurrence risk (*λ*_s_ = 4.5) in a UK study demonstrated familial aggregation and confirms the heritable component of DD.^9^ Two previous GWASs in ∼8,000 cases and ∼13,000 controls from three European countries identified 24 loci that contribute to DD risk.^10,11^ The proportion of heritability explained by the 24 genome-wide significant risk loci was estimated to be only 11.3%, indicating that additional genetic factors remain to be identified.^11^

To identify additional susceptibility loci and to further elucidate disease pathogenesis, we conducted GWASs in the UK Biobank resource and the FinnGen study and undertook a meta-analysis of these two datasets comprising 13,462 cases and 600,781 population controls. Furthermore, to identify putative causal variants and effector genes underlying GWAS signals, we determined colocalization of genome-wide significant hits with expression quantitative trait loci (eQTL). In addition, we evaluated whether genes under LD peaks of the top-associated GWAS variants were causative for human Mendelian diseases or animal-model phenotypes with a fibrotic component, defects in ECM homeostasis or bone development. Findings described in this study improve our understanding of the underlying molecular mechanisms of the DD pathogenesis and provide a rich resource that can facilitate the development of therapeutic interventions for DD and possibly for other forms of fibrotic diseases.

## Subjects and Methods

### UK Biobank cohort

UK Biobank is a prospective, population-based cohort study of over 500,000 participants enrolled at ages 40 to 69. The study participants were recruited between 2006 and 2010 at 22 assessment centers across the United Kingdom. Extensive amounts of phenotype and health-related data are available on each participant, including various biological measurements, indicators of lifestyle, biomarkers in blood and urine, multi-modal imaging of brain and body, and genetic data of various resolutions. Furthermore, information about health outcomes were obtained through linkage to electronic health and medical records that include inpatient hospital and primary care records, and data from cancer and death registries. More information about UK Biobank is available on the project’s website (see Web resources) and described elsewhere.^12^ Primary care data, which contain longitudinal clinical and prescription records obtained from general practices (GP), were available from ∼230,000 (∼45%) participants. Furthermore, the current GWAS was restricted to 409,559 (221,345 females and 188,214 males) Caucasians who self-identified as “white British” and that have a very similar genetic ancestry based on a principal component analysis (PCA) of the genotypes as described previously by Bycroft et al.^13^

### FinnGen study cohort

The FinnGen study is a population-based cohort study launched in 2017 as a public-private partnership aiming to identify genotype-phenotype relationships in the Finnish founder population.^14^ Health outcomes of study participants are followed up by linking to the national health registries since 1969. The present study was conducted using data from 342,499 individuals (190,879 females and 151,620 males) included in data release 8 (Fall 2021). Population outliers were excluded, and GWAS was performed in individuals with Finnish ancestry as determined by PCA.

### Ethics statement

Participants in the UK Biobank study provided signed electronic consents during assessment visits for follow-up through linkage to their health-related records. Ethics approval for the UK Biobank study was obtained from the North West Centre for Research Ethics Committee (11/NW/0382). Participants in FinnGen signed informed consent for biobank research in accordance with the Finnish Biobank Act (law 688/2012). Alternatively, separate research cohorts, collected prior the Finnish Biobank Act came into effect (in September 2013) and start of FinnGen (August 2017), were collected based on study-specific consents and later transferred to the Finnish biobanks after approval by Fimea (Finnish Medicines Agency), the National Supervisory Authority for Welfare and Health. Recruitment protocols followed the biobank protocols approved by Fimea. The Coordinating Ethics Committee of the Hospital District of Helsinki and Uusimaa (HUS) statement number for the FinnGen study is Nr HUS/990/2017. The FinnGen study is approved by Finnish Institute for Health and Welfare (permit numbers: THL/2031/6.02.00/2017, THL/1101/5.05.00/2017, THL/341/6.02.00/2018, THL/2222/6.02.00/2018, THL/283/6.02.00/2019, THL/1721/5.05.00/2019 and THL/1524/5.05.00/2020), Digital and Population Data Services Agency (permit numbers: VRK43431/2017-3, VRK/6909/2018-3, VRK/4415/2019-3), the Social Insurance Institution (permit numbers: KELA 58/522/2017, KELA 131/522/2018, KELA 70/522/2019, KELA 98/522/2019, KELA 134/522/2019, KELA 138/522/2019, KELA 2/522/2020, KELA 16/522/2020), Findata (permit numbers THL/2364/14.02/2020, THL/4055/14.06.00/2020, THL/3433/14.06.00/2020, THL/4432/14.06/2020, THL/5189/14.06/2020, THL/5894/14.06.00/2020, THL/6619/14.06.00/2020, THL/209/14.06.00/2021, THL/688/14.06.00/2021, THL/1284/14.06.00/2021, THL/1965/14.06.00/2021, THL/5546/14.02.00/2020, THL/2658/14.06.00/2021, THL/4235/14.06.00/2021) and Statistics Finland (permit numbers: TK-53-1041-17 and TK/143/07.03.00/2020 [earlier TK-53-90-20] TK/1735/07.03.00/2021). The Biobank Access Decisions for FinnGen samples and data utilized in FinnGen Data Freeze 8 include: THL Biobank BB2017_55, BB2017_111, BB2018_19, BB_2018_34, BB_2018_67, BB2018_71, BB2019_7, BB2019_8, BB2019_26, BB2020_1, Finnish Red Cross Blood Service Biobank 7.12.2017, Helsinki Biobank HUS/359/2017, Auria Biobank AB17-5154 and amendment #1 (August 17 2020), AB20-5926 and amendment #1 (April 23 2020), Biobank Borealis of Northern Finland_2017_1013, Biobank of Eastern Finland 1186/2018 and amendment 22 § /2020, Finnish Clinical Biobank Tampere MH0004 and amendments (21.02.2020 & 06.10.2020), Central Finland Biobank 1-2017, and Terveystalo Biobank STB 2018001.

### Phenotype definition

Case subjects in the UK Biobank were defined using multiple sources listed in **Table S1**. These sources include: 1) self-reported disease status obtained through a verbal interview by a trained nurse during the initial assessment visit; 2) physician-reported diagnostic codes recorded in inpatient hospital records according to the World Health Organization’s International Statistical Classification of Diseases and Related Health Problems (ICD) and 3) primary care data obtained from GP. From these three sources, a total of 8,846 Individuals were combined into the case group, among whom 31.2% were females and 68.8% were males. To minimize the effects of diagnostic misclassification, individuals with diagnosis records for various soft tissue disorders, including disorders of muscles (ICD10: M60-M63), disorders of synovium and tendon (ICD10: M65-M68) and other soft tissue disorders (ICD10: M70-M79) were excluded from the control group (n = 51,973). This resulted in a final control group of 347,659 white-British individuals. Over half of DD cases have undergone corrective surgeries of palmar fascia. The frequency of fasciectomies, or related hand surgeries in this cohort is in line with the reported percentage of fasciectomies in DD population, validating the ascertainment of case subjects from electronic health records. Cases in FinnGen were defined similarly using the ICD codes extracted from national health registries in Finland (**Table S1**). Individuals (n = 89,377) with soft tissue disorders were excluded from the control group. This resulted in a case and control groups of 4,616 and 253,122 individuals, respectively. Consistent with sex breakdown in the UK Biobank, among the Finnish cases 25.3% were females and 74.5% were males.

### Genotyping and imputation

Genotyping of the UK Biobank cohort was carried out using two closely related Affymetrix arrays, referred to as the UK BiLEVE and the UK Biobank Axiom arrays, both developed by Thermo Fisher Scientific. A subset of 49,950 participants were genotyped in 11 batches using the UKBiLEVE array (807,411 markers) and 438,427 participants were genotyped in 95 batches using the Axiom array (825,927 markers). These arrays share ∼95% of their marker content. Following genotype calling and initial data quality control (QC) by the vendor (i.e. Affymetrix), additional marker- and sample-level QC, phasing and imputation were carried out centrally by the UK Biobank as previously described by Bycroft et al.^13^ QC procedures were specifically chosen to account for large sample size, population structure and batch-based genotype calling. In brief, samples with over 5% genotype missingness and excess heterozygosity were removed. Markers with greater than 5% missing rate and a minor allele frequency (MAF) less than 0.0001 were excluded from imputation. Phasing was carried out using the SHAPEIT3 program.^15^ Using the IMPUTE4^16^ program and a combination of haplotype reference panels from the Haplotype Reference Consortium, UK10K and 1000 Genomes Project, a total of ∼97 million variants were imputed in 487,442 participants. We conducted genome-wide association testing in the third release version of the imputed genotype data. Samples collected for the FinnGen study were also genotyped using two custom designed Affymetrix (Thermo Fisher Scientific) arrays called FinnGen1 (657,675 markers) and FinnGen2 (664,510 markers). Legacy samples were genotyped with a combination of Illumina (Illumina) and Affymetrix arrays. Genotype imputation was performed with Beagle^17^ (v4.1) by using a population-specific reference panel of >20M variants, built from 8,554 high-coverage (25-30x) whole-genome sequences in Finns. Individuals with sex discrepancy, high genotype missingness (> 5%), excess heterozygosity (> 4 standard deviations from the mean), or non-Finnish ancestry were excluded. Variants with high missingness (> 2%), in Hardy-Weinberg disequilibrium (p < 1 × 10^-6^), or with minor allele count (MAC) of less than 3 were excluded.

### Association analysis

Single variant association between imputed genetic variation and the phenotype was tested using the mixed-effects logistic regression method implemented in the SAIGE R package.^18^ The UK Biobank cohort was analyzed with version 0.44.6.5 and the FinnGen cohort using the docker image v.0.36.3.2.fg of SAIGE. Variants with minor allele frequency (MAF) < 0.0001 and imputation INFO score < 0.8 were excluded from the analyses. After filtering, a total of 19,462,594 variants were tested in the UK Biobank cohort, and 17,510,894 variants were tested in the FinnGen cohort. Sex as inferred from genotypes, age at the time of recruitment (or at last follow-up in the FinnGen cohort), genotyping batch, and top 10 genotype-derived principal components (PCs) were used as covariates. Because of the nature of biobank-based studies, many participants tend to have biologically related individuals within the same cohort. For example, approximately ∼1/3 of the UK Biobank participants have at least one 1^st^, 2^nd^, or 3^rd^ degree relatives in the sample.^13^ We adjusted for within-sample relatedness, and for any residual population stratification, using a genetic relationship matrix (GRM) in the mixed-effects model. GRM was constructed using a linkage disequilibrium (LD)-pruned set of 83,967 variants, that were extracted from the BGEN files released by UK Biobank and selected from among 614,616 directly genotyped autosomal markers after excluding multiallelic variants, variants below MAF of 0.01 and those with imputation INFO scores less than 1. Pruning was performed using windows of 1Mb, a step-size of 10 markers and pairwise *r*^2^ < 0.05 with the PLINK2 software (v.2.00a2.3LM)^19^, using the command --*indep-pairwise* 1000 10 0.05. Top 10 PCs were calculated from an LD-pruned set of 83,967 variants using flashPCA.^20^ In the FinnGen cohort we used 55,139 LD-pruned variants for GRM calculation. These variants were selected from variants imputed with INFO score > 0.95 in all genotyping batches. Variants with > 3% missing genotypes were excluded as well as variants with MAF < 0.01. The remaining variants were LD pruned with a 1Mb window and *r*^2^ threshold of 0.1. Finally, saddle-point approximation (SPA) was applied to calibrate unbalanced case-control ratios in score tests based on logistic mixed models. The SAIGE method implements a logistic mixed-model approach and a saddle-point approximation (SPA) to the null distribution of the test statistic. Manhattan plots were generated using the *ggplot2* R package.^21^ Regional association plots were generated using the stand-alone version of LocusZoom (v1.3),^22^ using population specific LD-matrices.

### Meta-analysis

Prior to the meta-analysis, we first converted chromosomal coordinates of the FinnGen cohort summary statistics from the GRCh38/hg38 genomic build to GRCh37/hg19 using liftOver with the chain file hg38ToHg19.over.chain.gz obtained from the UCSC Genome Browser (see Web Resources). During liftover 61,663 variants (0.35%) failed to map to hg19 coordinates, among which only 5 variants had reached genome-wide significant association (p ≤ 5 × 10^-8^). These 5 variants belong to haplotypes with other genome-wide significant variants that were lifted over to their correct positions. Another 5,947 variants that cross-mapped to different chromosomes were excluded from the meta-analysis. None of these variants had reached genome-wide or suggestive level of evidence, and only five had p ≤ 0.001. Next, we standardized and QC’ed the summary statistics outputs using the *MungeSumstats* R package.^23^ Meta-analysis of the UK Biobank and FinnGen datasets was performed based on the inverse-variance weighted fixed effects model using the METAL software (version 2020-05-05).^24^

### Conditional SNP analysis

To identify independently associated variants within highly associated regions, we performed conditional and joint SNP analysis using the stepwise model selection procedure GCTA-COJO (command *--cojo-slct*) implemented in the GCTA software (v1.93.3beta).^25,26^ Reference LD structure was estimated using imputed genotypes of 10,000 randomly selected white-British individuals from the UK Biobank cohort. The distance assumed for complete linkage equilibrium was set to 10 Mb and a cut-off value of R^2^ = 0.9 was used to check for collinearity between the selected SNPs and those to be tested. Alleles with a frequency difference of over 0.2 between the reference sample and summary statistics were excluded. We nominated independent lead variants as from a single locus if they were within 250 kb of each other.

### Variant function analysis

Variants in LD with genome-wide significant SNPs were identified using the *LDlinkR* R package^27^ with LD data from the 1000 Genomes Project in the LDlink platform^28^ and annotated using the Variant Effect Predictor program (v104)^29^ in genome build GRCh37. VEP determines whether the genetic variant is in an exonic, intronic, or intergenic region and the functional consequence of the variant in different expression isoforms.

### eQTL colocalization

We selected 70 non-overlapping regions with a lead variant that was at least genome-wide significant (p < 5 × 10^-8^). For each of these regions, we sought to identify expression quantitative trait loci (eQTL) with a shared causal variant. For this, we used genome-wide summary statistics for eQTL calculated within a +/- 1Mb *cis*-window around gene transcription start sites from the GTEx project (v8; downloaded March 2020).^30^ This analysis included eQTL detected separately for 49 tissue types obtained from 838 donors. For each region associated with DD, we identified the lead variant and extracted summary statistics for all gene-tissue combinations in GTEx that had been tested for that variant. For each gene, the entire 2 Mb *cis*-candidate window tested in GTEx along with overlapping variants tested for association with DD was taken forward for colocalization analysis. After merging summary statistics for eQTL with DD GWAS meta-analysis summary statistics, we ran the COLOC method^31^ with default priors to identify gene-tissue pairs that share a causal variant affecting both gene expression and DD risk. In total, 2360 distinct genes and 76,321 gene-tissue pairs were tested for colocalization.

### LD Score Regression

SNP-based heritability (*h*^2^_SNP_) analysis was carried out using LD score regression under default settings using precalculated SNP LD scores from the HapMap 3 CEU population (https://data.broadinstitute.org/alkesgroup/LDSCORE/eur_w_ld_chr.tar.bz2) and provided with the LDSC (LD Score) software (v1.0.1).^32^ Heritability estimates were converted to a liability scale using the DD population prevalence of 0.05 to 0.25 and a combined sample prevalence of 0.022 (13,462/614,243) from both cohorts. Heritability was calculated using HapMap3 variants only.

### Gene set enrichment analysis

To test for overrepresentation of biological functions among genes in GWAS loci, we first positionally mapped genes within genome-wide significant loci to the nearest protein-coding genes using the SNP2GENE function of the FUMA webserver (v1.3.7).^33^ Next, using hypergeometric tests implemented in the GENE2FUNC function, we tested the prioritized list of genes against a collection of gene sets obtained from the Molecular Signatures Database (MsigDB)^34,35^ and from WikiPathways.^36^ MsigDB gene sets include hallmark, positional, curated, motif, computational, GO, oncogenic and immunologic signatures. As the background genes, against which the prioritized set of genes were tested, we used 19,142 protein-coding genes with unique entrez IDs. Correction for multiple hypothesis testing was performed using the Benjamini-Hochberg false discovery rate (FDR) per data source of tested gene sets. Gene sets with FDR-adjusted p ≤ 0.05 and the number of genes that overlap with the gene set > 1 were considered statistically significant. In addition, we performed gene-set analysis using the MAGMA^37^ (v1.06) test implemented in FUMA. In total, 4,761 curated gene sets and 5,917 GO terms obtained from MsigDB (v6.2) were included. Bonferroni correction was applied to all tested gene sets.

## Results

### Genome-wide association studies of DD in the UK and Finnish populations

Using a logistic mixed-effects model approach we tested ∼19.5 million imputed variants for association with DD in 8,846 cases and 347,659 population controls from white-British UK Biobank participants with European ancestry and ∼17.5 million imputed variants in 4,616 cases and 253,122 controls of Finnish ancestry from the FinnGen study. A total of 5,312 (**Figures S1A**) and 2,187 (**Figures S1B**) variants reached genome-wide significance (p ≤ 5 × 10^-8^) in each of these cohorts, respectively. These variants are shown in **Tables S2** and **S3**.

### Meta-analysis of GWAS identifies 83 risk loci

Since twenty-four genome-wide significant loci have already been associated with DD in a previous GWAS, setting a benchmark for disease risk loci, we pursued a joint analysis approach rather than the traditional two-stage GWAS. Such an approach has been suggested to offer better overall statistical power.^38^ Therefore, to maximize locus discovery, following association tests in these two distinct European population cohorts, we performed an inverse-variance weighted, fixed-effects meta-analysis. We identified 131 independent genome-wide significant signals that map to 83 unique loci (**Table S4**). Detailed summary statistics of all variants that reached genome-wide significance can be found in **Table S5**.

The genomic inflation factor (*λ*_GC_) of the meta-analysis association statistics was 1.14, whereas the LD score regression intercept was 1.04 (se = 0.01), indicating that most of the inflation in genome-wide association signals is due to true additive polygenic effects rather than a confounding bias, such as cryptic relatedness or population stratification. In line with these observations, we replicated association at all 24 known risk loci (26 independent hits) from the previous GWAS^11^; however, association at 59 loci were seen for the first time in our study, considerably increasing the number of disease susceptibility loci from the previously reported 24 to 83 loci (**Figure 1** and **Table 1**). Using LD score regression, we estimated the liability-scale SNP heritability (*h*^2^_SNP_) of DD to range between 25.18% (se = 0.03) and 41.32% (se = 0.05), given the sample proportion of cases equal to 0.02 and assuming that the disease prevalence ranges between 5% to 25% in the UK and Finnish populations.^3^

**Figure 1.**
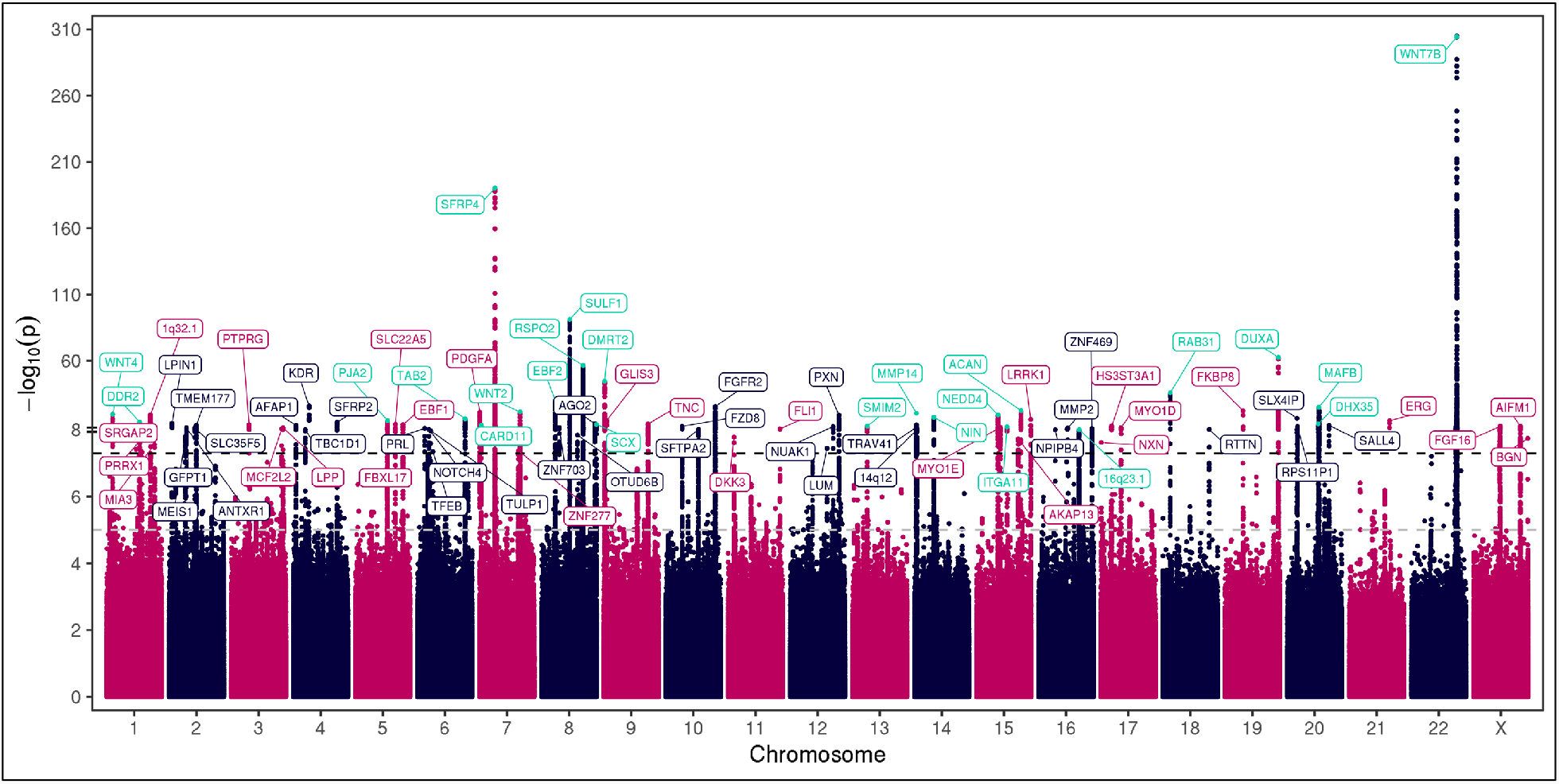
Manhattan plot showing results from meta-analysis of the GWAS of DD in UK Biobank and FinnGen. Negative log_10_ p values are plotted for each variant on the y-axis against their chromosomal position on the x-axis. Results are shown for a fixed-effect meta-analysis of effect estimates from GWASs in the UK Biobank and FinnGen cohorts, including a total of 13,462 cases and 600,781 controls. Eighty-three loci reached genome-wide significance and are labeled with the nearest or biologically most plausible candidate gene. Twenty-four known hits from the previous GWAS are colored in cyan. The threshold for genome-wide significance (p = 5 × 10^-8^) is indicated by a black dashed line, whereas the threshold for suggestive evidence of association (p = 1 × 10^-5^) is marked with a grey dashed line.

**Table 1:**
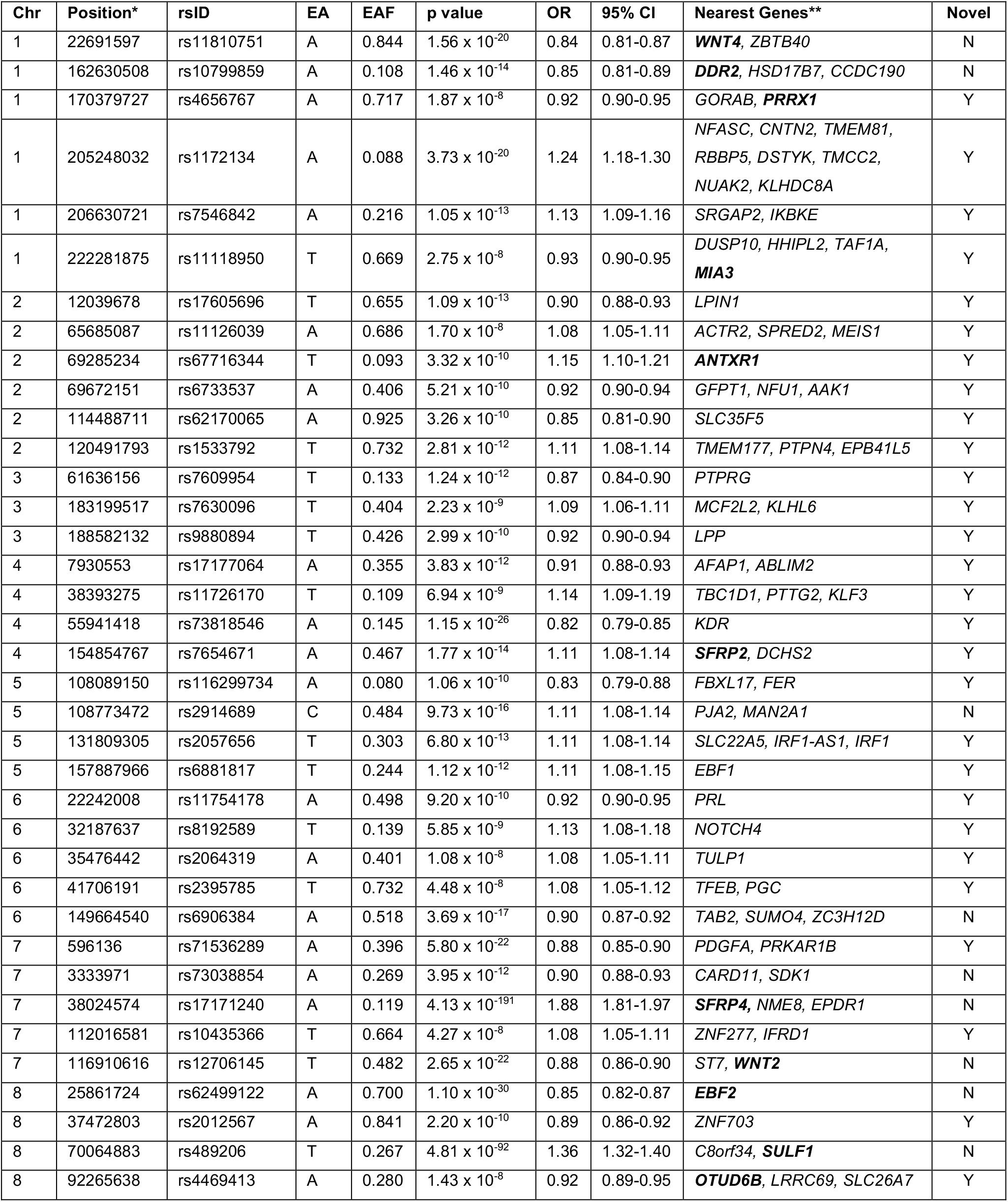
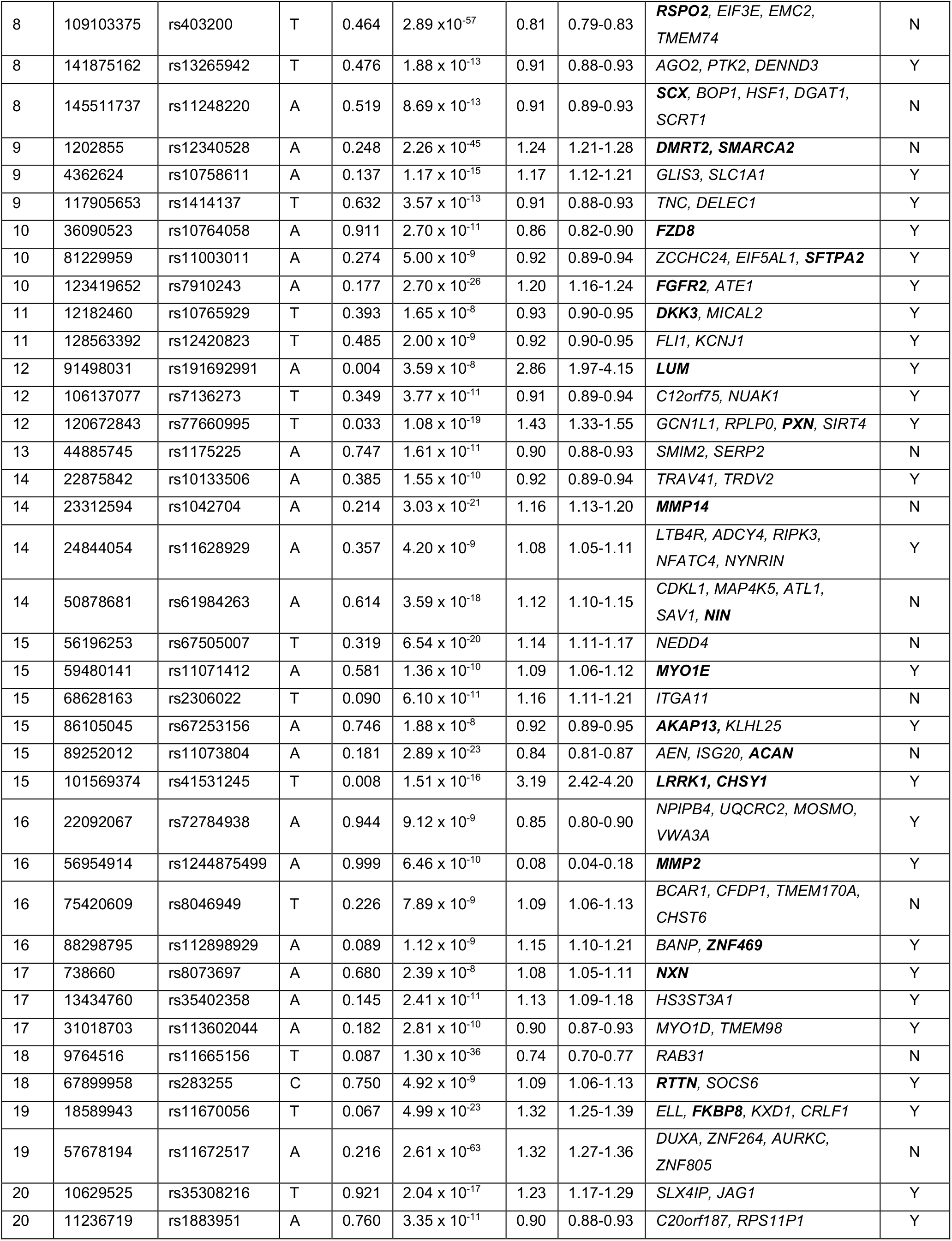
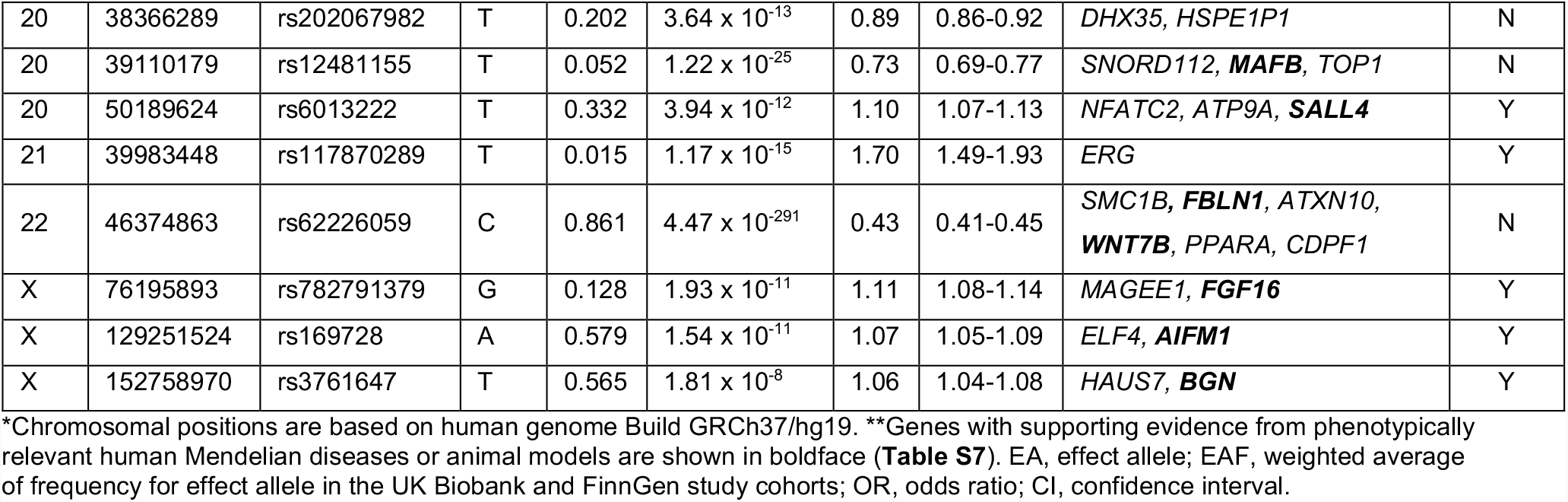
Lead variants at 83 loci associated with DD in the meta-analysis of GWAS in the UK Biobank and FinnGen cohorts (p ≤ 5 × 10^-8^).

### Variant annotation and identification of candidate causal genes

Using the VEP tool^29^ we annotated all 9,213 genome-wide significant variants from the meta-analysis and variants in strong LD (*r*^2^ ≥ 0.8 in EUR from the 1000 Genomes Project) with independent lead SNPs that were excluded or that did not reach genome-wide significance due to low allele frequency. We found that 325 variants reside in coding regions, 48 of which result in amino acid changes in 24 genes (**Table S6)**. In addition, we identified one splice-site donor (rs58264281 in the 5’UTR exon of *AURKC*), one stop-gained (rs76807453 in *TRGV11*) and one in-frame deletion variant (rs370403371 in isoform ENST00000518442.5 of *EIF3E*). The remaining variants fall into UTRs, intronic or intergenic regions, suggesting that these variants might modulate disease risk through mRNA instability or expression changes of their target genes. *TRGV11* is a poorly characterized gene and the causal gene in that locus is likely the previously reported, and experimentally validated Wnt antagonist gene *SFRP4*.^11^ Similarly, the in-frame deletion is likely an LD variant of the true causal variant linked to the previously reported Wnt signaling pathway gene, *RSPO2*. It has yet to be determined whether rs58264281 has any impact on the function of *AURKC* or causally implicated in DD. One of the non-synonymous variants (p.Asp273Asn, rs1042704, OR = 1.16, p = 3.03 × 10^-21^) is a replication of a previously reported etiological change in the membrane type 1 matrix metalloproteinase (MT1-MMP) protein, which has been shown to reduce the collagenolytic activity of the enzyme.^39^

### Finnish-enriched protein coding risk variants

Compared to other populations, Finns carry a significant excess of rare and low-frequency deleterious variants.^40^ We identified three novel associations with such Finnish-enriched and non-synonymous variants. The association signal on chromosome 16q12.2 is likely driven by the rare, high effect missense variant p.Ser396Arg (rs200772153, OR = 6.42, p = 9.13 × 10^-8^) in the *MMP2* gene, coding for the matrix metallopeptidase 2 protein. There are 5 SNPs spread across a 643-kb region that reach genome-wide significant threshold at this locus, including the lead SNP rs1244875499 with a p-value of 6.49 × 10^-10^. The rare missense variant, which is in moderate LD with the lead SNP (*r*^2^ = 0.6), is almost nonexistent outside Finland, while it has an allele frequency of 0.14% in the Finnish population, likely due to a founder effect. Moreover, according to both PolyPhen-2^41^ and SIFT^42^, the risk allele is predicted to impair the structure or function of the enzyme. *MMP2* is an attractive candidate gene, because protein truncating mutations in *MMP2* have been shown to cause a recessive form of the ‘vanishing bone’ syndrome, manifested by multicentric osteolysis with carpal and tarsal resorption, severe arthritic changes, osteoporosis, palmar and plantar subcutaneous nodules, and distinctive facies^43^ (MIM: 259600).

The lead SNPs at the other two loci are the Finnish-enriched non-synonymous variants, p.Thr967Met (rs41531245, OR = 3.18, p = 1.51 × 10^-16^) in *LRRK1* and p.Arg310Cys in *LUM* (rs191692991, OR = 2.86, p = 3.59 × 10^-8^) (**Figure 2**). Finnish enrichment here is calculated as AF_(FIN)_ / AF_(NFSEE)_ in the gnomAD 2.1.1 database (see Web Resources), where NFSEE stands for non-Finnish-non-Swedish-non-Estonian European. The ancestral, non-risk amino acid residues of both variants are highly conserved across many species, and the risk-increasing alleles are predicted as probably damaging by PolyPhen-2. Notably, the association signals of both variants were driven entirely by the Finnish cohort, as the frequencies of these variants were extremely low in the UK Biobank, and compared to non-Finnish Europeans these two variants are enriched in Finns by 15- and 34-fold, respectively.^44^ Interestingly, we also identified an independent signal 293 Kb downstream of *LRRK1* transcription start site, that yields genome-wide significant association (rs4965806, OR = 0.92, p = 5.73 × 10^-9^) in the meta-analysis. Due to the proximity of lead variants, we assigned these signals to a single locus. However, these associations seem to be driven by separate genes. The second signal covers the entire chondroitin sulfate synthase 1 gene, *CHSY1*, which has been linked to a rare, syndromic recessive preaxial brachydactyly.^45^ *LRRK1* encodes the leucine-rich repeat serine/threonine-protein kinase 1, which is thought to play a role in bone mass regulation. *Lrrk1*-null mice develop severe osteopetrosis, reduced bone resorption in endocortical and trabecular regions, and increased bone mineralization.^46^ Loss-of-function variants in this gene have been linked to the rare disorder osteosclerotic metaphyseal dysplasia (MIM: 615198).^47,48^ *LUM* on the other hand codes for an extracellular matrix protein called lumican, a small leucine-rich repeat proteoglycan (SLRP) that regulates the assembly of collagens into higher-order fibrils in connective tissues. Due to deregulated growth of abnormally thick collagen fibrils, mice homozygous for a lumican null mutation develop Ehlers-Danlos syndrome (EDS)-like skin fragility and laxity, and bilateral corneal opacification.^49^

**Figure 2:**
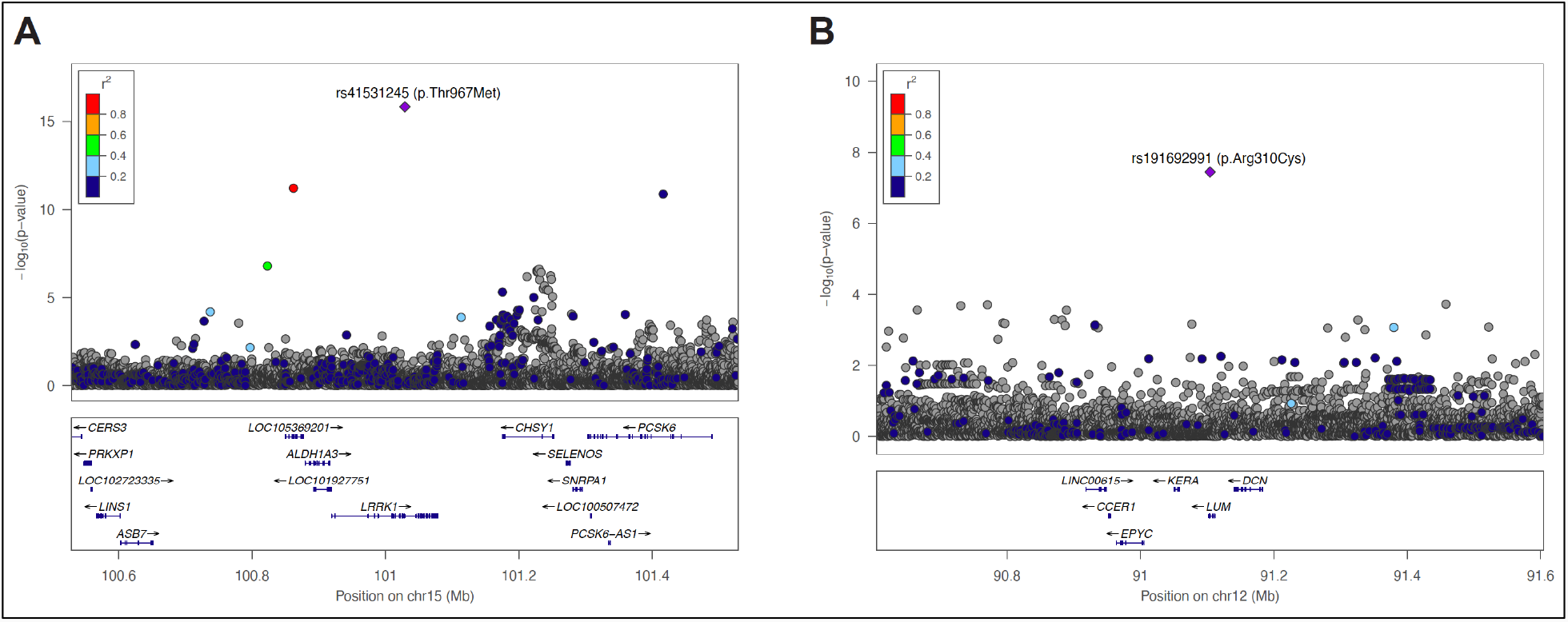
LocusZoom plots of the Finnish-enriched missense variants in the A) *LRRK1* and B) *LUM* genes. Negative log_10_ p values are shown for variants within a 1Mb region centered at the reference SNP labeled with a purple diamond. Pairwise LD (*r*^2^) between the reference SNP and other variants in the region are indicated by color. The *r*^2^ values were estimated from whole-genome sequencing data of 8,554 Finns. Genomic coordinates are based on the human genome build GRCh38/hg38.

### Genes involved in fibrosis

To gain insights into the potential biological mechanisms underlying DD pathogenesis, we conducted a series of *in-silico* functional analyses using various bioinformatics tools and publicly available datasets. We first queried OMIM for rare, monogenic human diseases and the MARRVEL^50^ and Alliance of Genome Resources^51^ databases for relevant animal model phenotypes for association with genes that were located either directly under the LD peaks or flanking the risk haplotypes when the signal fell into an intergenic region. Defects in genes at over 30 loci were found to cause a variety of phenotypes with profound fibrotic component, and that predominantly involve bone or connective tissue development and maintenance (**Table S7**). For example, we identified association with an 81 kb-long haplotype that covers the entire intergenic region between *SFTPA2* and *ZCCHC24*, and that contains the *EIF5AL1* gene, with the lead SNP (rs11003011, OR = 0.92, p = 5 × 10^-9^) located approximately 90 kb downstream of the *SFTPA2* gene. *SFTPA2* encodes the surfactant protein A2 mutated in a rare, familial form of IPF.^52^ The lead SNP is in nearly complete LD with the rs7920426 (*r*^2^ = 0.97, p = 1.52 × 10^-8^) SNP located in a candidate *cis*-regulatory element, identified and classified by the ENCODE project^53^ using various biochemical signatures, in this intergenic region. It is however unknown whether this region regulates the expression of the *SFTPA2* gene.

Furthermore, we replicated association at the *NEDD4* gene locus (rs67505007, OR = 1.14, p = 6.54 × 10^-20^), whose murine homolog *Nedd4l* has been implicated in lung fibrosis.^54,55^. The lead SNP is in complete LD with the intronic variant rs8032158 (*r*^2^ = 1.0, p = 7.61 × 10^-20^) that has been associated with predisposition to skin keloid formation, a dermal fibroproliferative growth caused by pathologic wound healing, in the Japanese population.^56^

These two genes were not the only genes with links to other fibrotic diseases. The lead SNP rs67253156 (OR = 0.92, p = 1.88 × 10^-8^) at one of the novel loci is strongly correlated (*r*^2^ = 0.96) with a putative, deleterious missense variant (rs745191, p.Gly624Val) in *AKAP13*. The *AKAP13* gene region has been associated with IPF in European GWAS.^57,58^ The IPF risk-increasing allele was also found to increase the expression level of the *AKAP13* mRNA in lung tissue. *AKAP13* encodes the A-kinase anchoring protein 13, a Rho guanine nucleotide exchange factor regulating activation of RhoA, which is known to be involved in profibrotic signaling pathways.^59^ These findings suggest potential shared genetic risk between DD and pulmonary fibrosis, although the extent of pleiotropy between DD and pulmonary fibrosis has yet to be determined.

### eQTL colocalization with GWAS genes

Within 1 Mb windows in each direction from the lead SNP we identified approximately 1,430 protein-coding genes. Although LD-blocks are ideal to localize genomic regions carrying causal variants, in many instances such variants fall into long distance-acting *cis*-regulatory regions, such as transcriptional enhancers, that can extend well beyond disease-associated LD-blocks. To identify putative regulatory variants that likely affect expression of causal genes, we performed eQTL colocalization analysis.^31^ Of 76,321 tissue-gene pairs tested for colocalization, 228 pairs representing 81 distinct genes showed strong evidence of colocalization [Posterior Probability (PP) > 0.8] (**Table S8**). Skin, lung, whole blood, pancreas, tibial nerve, and fibroblasts were the tissues with the greatest number of colocalizations, while brain tissues tended to have the lowest number of colocalizations (**Figure S2**).

The lead SNP rs4656767 (OR = 0.92, p = 1.87 × 10^-8^) at the chromosome 1q24 signal is located in the intergenic region upstream of the *GORAB* gene. However, the DD association signal colocalizes with eQTL of the *PRRX1* gene in cultured fibroblast cells (PP = 0.97) and lung tissues (PP = 0.93) in the non-coding region with multiple, putative enhancer elements, suggesting that a regulatory variant might drive the association. PRRX1 (paired related homeobox 1), a member of the paired homeobox family, regulates the formation of preskeletal condensations from undifferentiated mesenchyme. As the genetic association with DD indicates, *PRRX1* is not only active during embryogenesis, but it seems to contribute to fibrotic processes in adult tissues. For example, fibroblasts expressing *Prrx1* were shown to drive acute and chronic fibroses in the ventral dermis during wound repair.^60^ Recently, *PRRX1* has been implicated in the pathogenesis of IPF, where the expression of the gene both at the transcriptional and protein levels was increased in affected compared to healthy lung tissues. Moreover, inhibition of *Prrx1* attenuated fibrotic remodeling *in vivo* in the bleomycin mouse model of lung fibrosis and *ex vivo* in precision-cut lung slices.^61^

### Gene set enrichment

To determine biological pathways enriched for genes within risk loci, we performed gene enrichment analysis using the FUMA web application.^33^ Positionally mapped genes prioritized within the 83 risk loci were assessed for their overrepresentation among a set of curated genes and pathways obtained from MsigDB and WikiPathways, respectively. Five pathways were statistically significantly enriched (FDR-adjusted p value < 0.05), including epithelial tube branching involved in lung morphogenesis, lung morphogenesis, cell substrate adhesion, response to growth factor and extracellular structure organization (**Figure S3**). Gene-set analysis using the MAGMA method, that takes into account the full distribution of SNP p values within genic regions, yielded similar results below the Bonferroni corrected p < 0.05, but also identified a GO term related to skeletal system development (**Table S9**). Taken together, these associations further support the predominant role of genes playing role in assembly and homeostasis of connective tissue and ECM in DD. At the same time, very low number of immune system genes in GWAS loci points toward a limited or no role of inflammatory processes in disease initiation. Crosstalk between the Wnt signaling pathway and inflammation was alluded as a potential etiologic factor in a previous GWAS, which also identified very few inflammatory genes.^11^ Lack of well-known inflammation drivers in the expanded genetic landscape of DD here confirms that disease predisposition is not driven by the immune system, but that inflammation observed in DD tissues is likely secondary to the fibrotic processes.

## Discussion

DD is a peculiar condition that affects a restricted area within the palmar fascia. Due to its high familial aggregation, earlier studies considered DD as a dominantly inherited disease with variable penetrance.^62^ However, recent GWASs including our study here demonstrate remarkable polygenicity for an ostensibly simple, tissue-restricted disease that affects only a limited area of the hand. To the best of our knowledge, our study is the largest genome-wide study that has also taken an in-depth examination of individual risk loci to explore the biological mechanisms underlying genetic etiology of the disease.

By aggregating data from two large nationwide biobanks, we identified 83 loci associated with susceptibility to DD, more than tripling the number of common variant associations to DD susceptibility. More importantly, we replicated association at all known DD-risk loci. At three loci the lead variants were the exact same lead variants (rs1042704, rs2306022 and rs11672517) reported by Ng et al. in the most recent GWAS.^11^ Two of these are non-synonymous changes in the *MMP14* and *ITGA11* genes. At remaining loci, the lead variants were in moderate-to-strong LD with previously identified lead variants, yet all reached genome-wide significance in our study (**Table S5**). For example, the lead SNP rs10799859 (p = 1.46 × 10^-14^) at the *DDR2* (discoidin domain receptor tyrosine kinase 2) gene locus was in moderate LD (*r*^2^ = 0.6 in EUR) with the previously reported lead SNP rs17433710 (p = 1.70 × 10^-10^). Both SNPs are located in the same intron of the *DDR2* gene, which is an attractive candidate causal gene in this locus as it has been implicated in fibrosis in numerous studies. Mice deficient for both *Dddr1* and *Dddr2* are resistant to bleomycin-induced lung fibrosis.^63,64^ Inhibition of discoidin domain receptors have been under investigation as a potential therapeutic approach for fibrosis, such as in IPF.^65^ Moreover, autosomal recessive mutations in *DDR2* cause the rare spondylo-meta-epiphyseal dysplasia (SMED, short limb-hand type [MIM 271665]),^66^ whereas autosomal dominant, activating mutations cause Warburg-Cinotti syndrome (WCS, [MIM: 618175])^67^ manifested by progressive corneal neovascularization, keloid formation, chronic skin ulcers, acro-osteolysis, joint contractures and palmar fibrotic bands. Overlapping features, such as the palmar fibrotic bands, prompted us to explore whether there were more genes within DD-risk loci that cause similar, rare monogenic limb abnormalities or play a role in early limb development in general. We thought that such connections can potentially provide clues to the unique, hand-specific manifestation of DD.

In previous GWASs, five out of twenty-four risk loci harbored genes coding for members of the Wnt signaling pathway or coding for their co-signaling molecules.^10,11^ Wnt proteins function as intercellular signals to regulate the proliferation of cells and confer polarity and asymmetry to cells giving shape to tissues. These signaling proteins are required in numerous contexts – initially in early development and later during the growth and maintenance of various adult organ tissues.^68^ Interestingly, nearly all Wnt ligand genes and genes for their receptors, called frizzled (Fzd), are expressed in different regions of developing limb buds at one point or another.^69^ The fundamental role of Wnt proteins is reflected in different developmental abnormalities caused by rare pathogenic mutations in these genes, several of which have particularly been linked to congenital limb malformations. At the cellular level, the biochemical signaling mechanisms of different Wnt proteins are very similar. However, the differences between Wnt-linked phenotypes can be attributed to discrete and unique spatiotemporal expression patterns of the Wnt genes. For example, loss-of-function mutations in *WNT3* cause tetra-amelia syndrome or complete absence of all four limbs^70^, whereas mutations in the *WNT10B* gene cause split-hand/foot malformation.^71^ Similarly, mutations in the *RSPO2* gene, a Wnt antagonist, cause tetra-amelia with a combination of other severe anomalies.^72^ Mutations in several other Wnt signaling component genes such as *ROR2, WNT5A, DVL1, DVL3, FZD2*, or *NXN* are linked to Robinow syndrome^73^, a congenital malformation characterized by limb shortening and short stature. Therefore, the genetic association of Wnt signaling pathway with a limb-specific disease such as DD is not surprising. Perhaps genetic variants that were evolutionarily selected for other physiological or anthropomorphic traits, such as height for example, exhibit deleterious effects later in life. Indeed, supporting this hypothesis, several DD-risk loci harbor genes (*WNT4, ACAN, PTPRG, FER*) that show significant association with adult human height in the GIANT consortium GWAS.^74^ Including several Wnt genes causative of rare limb malformation, we found at least 15 genes that result in monogenic, congenital limb malformation when mutated (**Table S7**).

In this study we discovered four additional loci that harbor genes coding for key members or interacting proteins of the Wnt signaling pathway, including *DKK3, SFRP2, NXN* and *FZD8. DKK3* codes for the Dickkopf-related protein 3, a member of the dickkopf family of Wnt inhibitors. In developing embryo limbs, *Dkk3* is detected in the area of the carpal condensations and in metacarpal-phalangeal joints.^75^ Metacarpal-phalangeal joints are typically the first and most severely affected digital joints in DD development. Secreted frizzled-related protein 2 (SFRP2) is a soluble modulator of Wnt signaling. Loss of *Sfrp2* cause limb defects in mice with mesomelic shortening and consistent shortening of all autopodal elements that clinically manifests as brachydactyly in humans.^76^ In addition, these animals display soft-tissue syndactyly of the hindlimb. The brachydactyly phenotype is caused by decreased chondrocyte proliferation and delayed differentiation in distal limb chondrogenic elements. The third gene *NXN* codes for nucleoredoxin, a redox-dependent negative regulator of the Wnt signaling pathway, which is also highly expressed in the developing limb buds of mice.^77^ Biallelic loss-of-function mutations in *NXN* cause the recessive form of Robinow syndrome with mesomelic shortening and finger brachydactyly and clinodactyly.^73^

Finally, *FZD8*, coding for one of the ten Frizzled class receptors for Wnt ligands, and the only receptor gene of the Wnt pathway that has been implicated in DD, is located downstream of the novel GWAS signal on chromosome 10p11.2. Although the risk haplotype at the *FZD8* gene locus falls within an intergenic region, there are two experimentally validated gene enhancers located upstream of the gene that likely regulate expression of the *FZD8* gene. The risk haplotype partially overlaps with the enhancer annotated as hs1589 in the VISTA Enhancer Browser,^78^ which is located within the intronic region of the long non-coding RNA gene *PCAT5*. This enhancer shows broad regulatory activity, whereas the second enhancer, hs1567, which does not seem to contain any known variants in LD with the risk-increasing variants, drives expression of the *LacZ* reporter gene specifically in the apical ectodermal ridge. It has yet to be determined whether these two enhancer elements interact with one another and act upon the *FZD8* gene to induce its expression in limb tissues. Through such a mechanism the DD-associated variants may lead to aberrant expression of the *FZD8* gene in adult limb tissue. Alternatively, there may be an unidentified copy-number variation tagged by the risk haplotype that could affect the expression level or dose of the *FZD8* gene. Nonetheless, the overrepresentation of the WNT signaling pathway genes in DD GWAS loci suggests that *FZD8* is a top candidate causal gene within the chr10p11.2 locus.

Furthermore, TGF-β is a well-establish mediator of tissue fibrosis. TGF-β-driven pro-fibrotic signaling has been shown to act through this particular Wnt receptor.^79^ Knock-out mouse model with complete deletion of the *Fzd8* gene are resistant to bleomycin-induced lung fibrosis^80^ demonstrating the therapeutic potential of inhibiting the FZD8 receptor activity not only in DD but in other fibrotic diseases, such as IPF, in general. Additionally, multiple lines of evidence demonstrate a key role of the Wnt signaling pathway in general in fibrosis and myofibroblast differentiation in multiple disease indications such as systemic sclerosis^81^ and IPF^82^. Overall, these results indicate that members of the Wnt pathway may provide key targets in modulating myofibroblast biology backed by human genetic evidence.

Another class of signaling factors and receptors that emerged in our study for the first time is of the fibroblast growth factors (FGF) signaling pathway. Interplay between the Wnt and FGF signaling pathways has been long known to play a key role at various stages of limb development.^83^ We identified two regions that harbor genes coding for the FGFR2 receptor (rs7910243, p = 2.70 × 10^-26^, OR=1.20), and FGF16 (rs782791379, p = 1.93 × 10^-11^, OR=1.11). FGFR2 is a well-characterized receptor with gain-of-function mutations causative of a range of skeletal abnormalities. These include abnormal bone development, craniofacial and skeletal synostosis or camptodactyly. Protein truncating mutations in *FGF16* cause X-linked, recessive metacarpal 4-5 fusion (MF4; MIM: 309630) characterized by partial or complete fusion of the 4^th^ and 5^th^ metacarpals.^84,85^ Strikingly, metacarpal-phalangeal joints of the 4^th^ and 5^th^ digits are also the most frequently involved sites in DD.

In conclusion, association of genes involved in fibrosis in different organ tissues, such as heart, lung, and likely others, with a single but common, complex disease attest to the fascinating feature of DD as a human model disease to uncover the causal mechanisms of pathologic fibrosis through genetics. The largest genetic study described here further contributes to our understanding of the genetic architecture of DD and provides a comprehensive list of genes and loci for experimental validation.

## Supporting information

FinnGen Author List

## Data Availability

All data produced in the present study are available upon reasonable request to the authors.

## Supplemental Data

Supplemental Data include three figures and nine tables.

## Acknowledgements

Data from the UK Biobank was obtained under the application number 26041. We would like to thank AbbVie employees Jason Grundstad and Mark Reppell for their assistance with data acquisition from the UK Biobank. UK Biobank was established by and receives core funding from the Wellcome Trust medical charity, Medical Research Council, Department of Health, Scottish Government, and the Northwest Regional Development Agency. It has also had funding from the Welsh Government, British Heart Foundation, Cancer Research UK, and Diabetes UK. UK Biobank is supported by the National Health Service (NHS). The FinnGen study is funded by two grants from Business Finland (HUS 4685/31/2016 and UH 4386/31/2016) and the following industry partners: AbbVie Inc., AstraZeneca UK Ltd, Biogen MA Inc., Bristol Myers Squibb (and Celgene Corporation & Celgene International II Sàrl), Genentech Inc., Merck Sharp & Dohme Corp, Pfizer Inc., GlaxoSmithKline Intellectual Property Development Ltd., Sanofi US Services Inc., Maze Therapeutics Inc., Janssen Biotech Inc, and Novartis AG. Following biobanks are acknowledged for delivering biobank samples to FinnGen: Auria Biobank (www.auria.fi/biopankki), THL Biobank (www.thl.fi/biobank), Helsinki Biobank (www.helsinginbiopankki.fi), Biobank Borealis of Northern Finland (https://www.ppshp.fi/Tutkimus-ja-opetus/Biopankki/Pages/Biobank-Borealis-briefly-in-English.aspx), Finnish Clinical Biobank Tampere (www.tays.fi/en-US/Research_and_development/Finnish_Clinical_Biobank_Tampere), Biobank of Eastern Finland (www.ita-suomenbiopankki.fi/en), Central Finland Biobank (www.ksshp.fi/fi-FI/Potilaalle/Biopankki), Finnish Red Cross Blood Service Biobank (www.veripalvelu.fi/verenluovutus/biopankkitoiminta) and Terveystalo Biobank (www.terveystalo.com/fi/Yritystietoa/Terveystalo-Biopankki/Biopankki/). All Finnish Biobanks are members of BBMRI.fi infrastructure (www.bbmri.fi). Finnish Biobank Cooperative -FINBB (https://finbb.fi/) is the coordinator of BBMRI-ERIC operations in Finland. The Finnish biobank data can be accessed through the Fingenious® services (https://site.fingenious.fi/en/) managed by FINBB. We would like to thank the clinical, analysis and administrative teams of the FinnGen study. Finally, we would also like to thank all participants of the UK Biobank and FinnGen studies for generously providing samples and access to their medical records.

## Web resources

OMIM, https://www.omim.org

MARRVEL, http://marrvel.org

Alliance of Genome Resources, https://www.alliancegenome.org

SAIGE, https://github.com/weizhouUMICH/SAIGE

Finngen/saige, https://hub.docker.com/r/finngen/saige

LDSC, https://github.com/bulik/ldsc

PLINK, https://www.cog-genomics.org/plink

METAL, https://genome.sph.umich.edu/wiki/METAL

LocusZoom, http://locuszoom.org

UCSC Genome Browser, https://genome.ucsc.edu

Vista Enhancer Browser, https://enhancer.lbl.gov

ENCODE Project, https://www.encodeproject.org

UK Biobank Data Showcase, https://biobank.ndph.ox.ac.uk/showcase

FinnGen study, https://www.finngen.fi

LDlink, https://ldlink.nci.nih.gov

gnomAD, https://gnomad.broadinstitute.org

FUMA GWAS, https://fuma.ctglab.nl

