## Supplemental Data for "A meta-analysis of genome-wide association studies in 614,243 individuals identifies 59 novel susceptibility loci underlying Dupuytren’s contracture"

**Supplemental Figures**

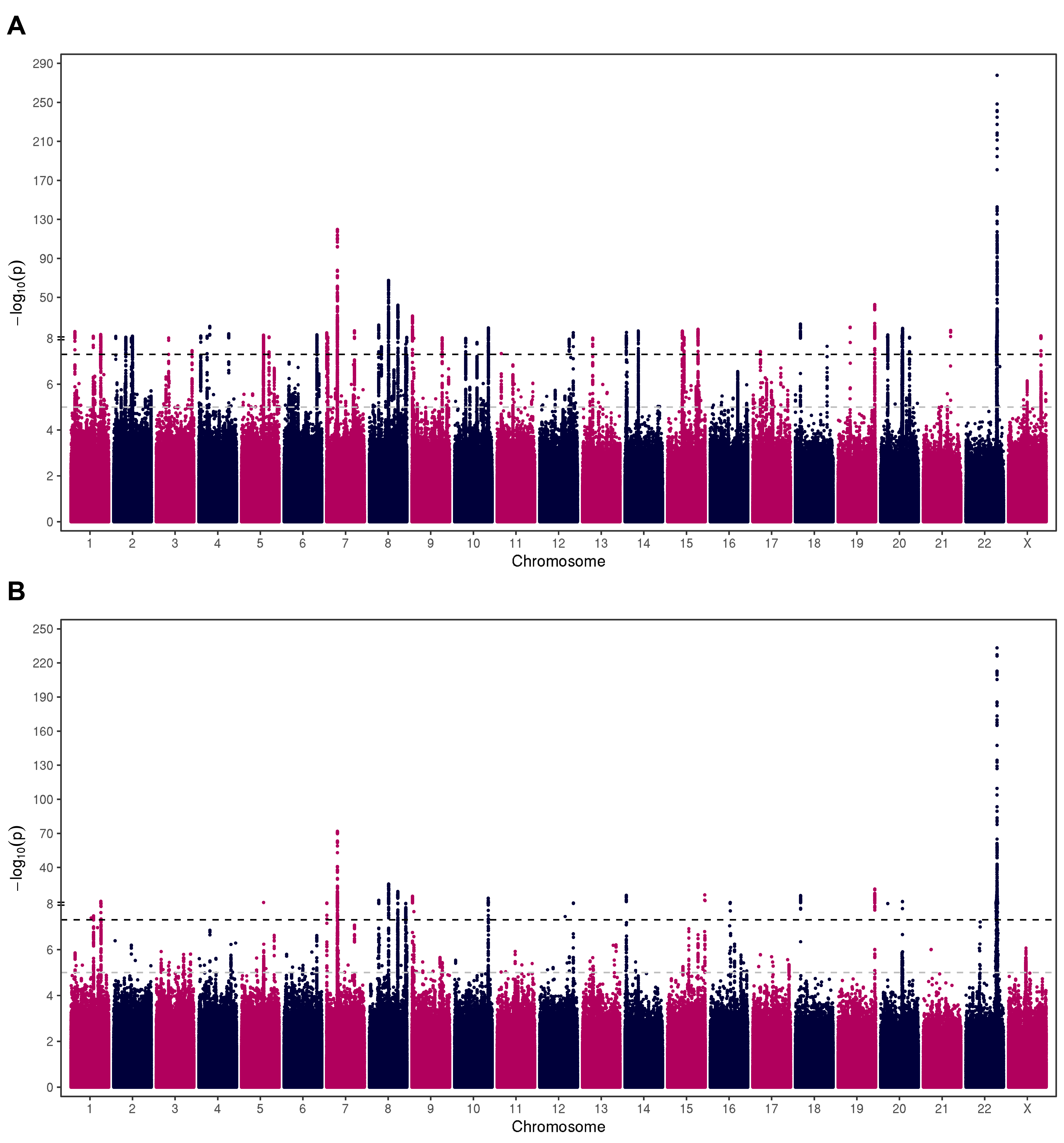

**Figure S1. Manhattan plots showing population specific GWAS results of A) 8,846 DD cases and 347,659 controls from the UK Biobank resource and B) 4,616 cases and 253,122 controls from the FinnGen study.** Negative log_10_ p values (y axis) are plotted for each variant against their chromosomal position (x axis). The threshold for genome-wide significance (p = 5 x 10^-8^) is indicated by a black dashed line, whereas the threshold for suggestive evidence of association (p = 1 x 10^-5^) is marked with a grey dashed line.

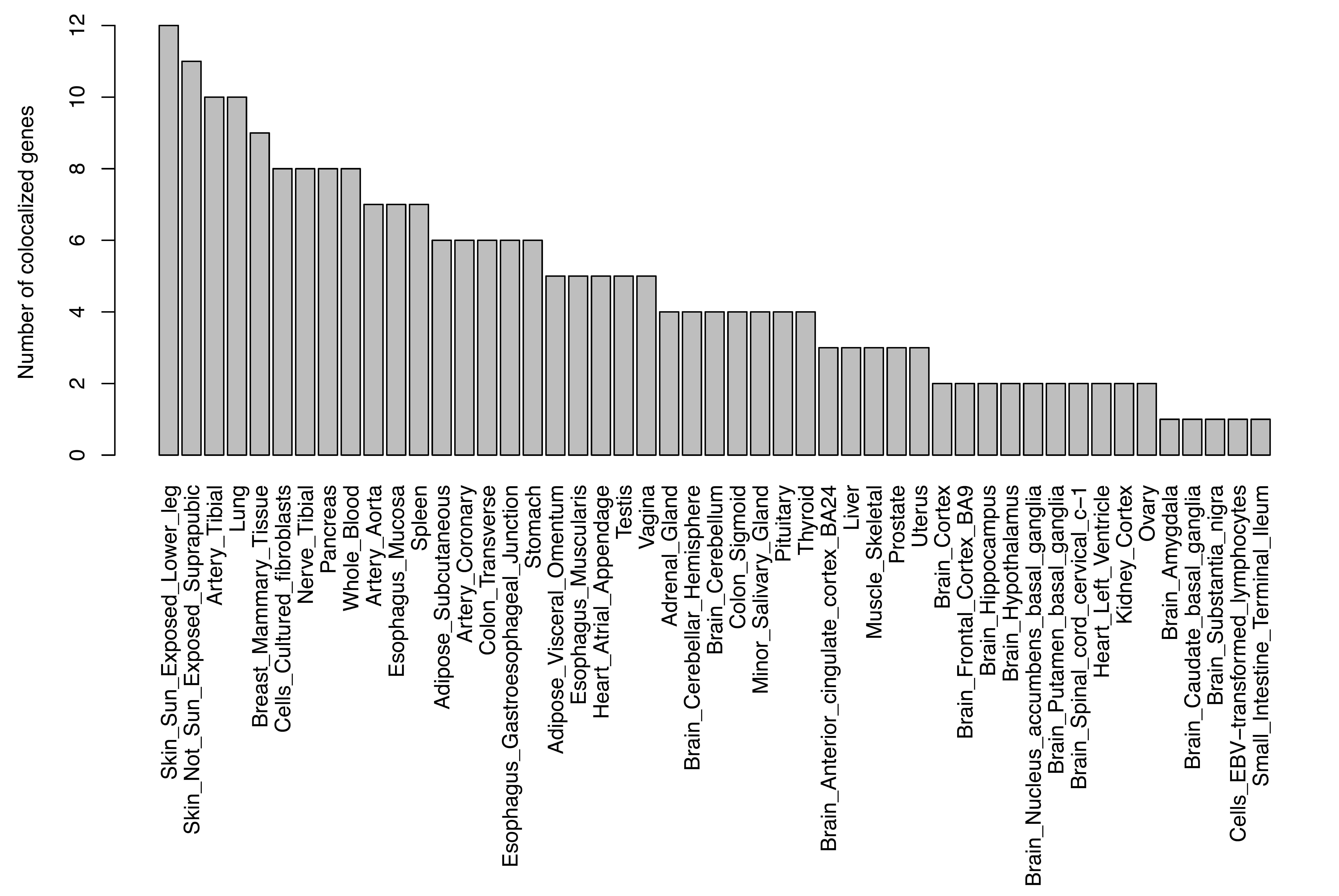

**Figure S2: eQTL colocalized genes**

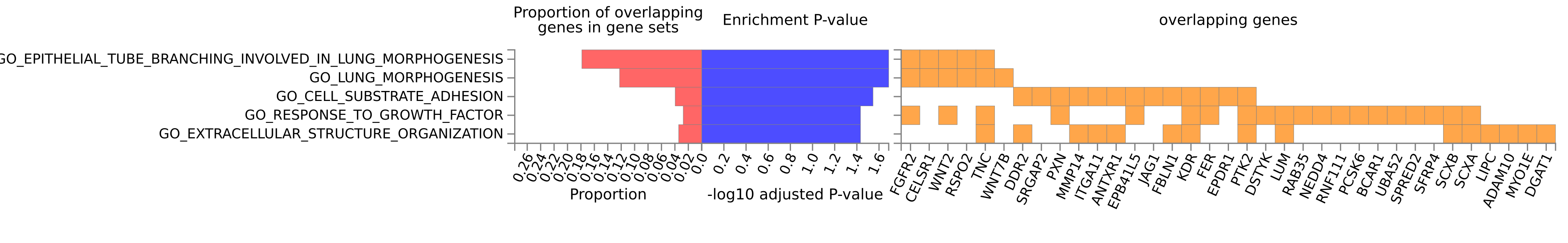

**Figure S3: FUMA Pathway analysis**

**Supplemental Tables**

**Table S1:** **Phenotype definitions**

**Table S2: Genome-wide significant variants in the UK Biobank cohort**

**Table S3: Genome-wide significant variants in the FinnGen study cohort**

**Table S4: Independent lead variants from conditional and joint SNP analysis (GCTA-COJO)**

**Table S5: Genome-wide significant variants from the meta-analysis of GWAS**

**Table S6: VEP output**

**Table S7: Rare human monogenic diseases and animal models associated with GWAS genes**

**Table S8: Significant genes from eQTL colocalization analysis**

**Table S9: MAGMA Gene-Set Analysis**
